# Multisystem Inflammatory Syndrome in Children Managed in the Outpatient Setting: An EHR-based cohort study from the RECOVER program

**DOI:** 10.1101/2022.08.25.22279225

**Authors:** Ravi Jhaveri, Ryan Webb, Hanieh Razzaghi, Julia Schuchard, Asuncion Mejias, Tellen D. Bennett, Pei-Ni Jone, Deepika Thacker, Grant S. Schulert, Colin Rogerson, Jonathan D. Cogen, L. Charles Bailey, Christopher B. Forrest, Grace M. Lee, Suchitra Rao, the RECOVER consortium

## Abstract

Using electronic health record data combined with primary chart review, we identified 7 children across 8 pediatric medical centers with a diagnosis of Multisystem Inflammatory Syndrome in Children (MIS-C) who were managed as outpatients. These findings should prompt a discussion about modifying the case definition to allow for such a possibility.

## Introduction

At the outset of the COVID-19 pandemic, an uncommon but severe manifestation of SARS-CoV-2 infection was described in children. While Initially labeled as an unusual “Kawasaki-like” syndrome, it subsequently became clear that many findings were inconsistent with Kawasaki Disease^1-4^. The condition was ultimately named “Multisystem Inflammatory Syndrome in Children” or MIS-C and included within the broad collection of post-acute sequelae of SARS-CoV-2 (PASC) conditions. The Centers for Disease Control and Prevention (CDC) created an MIS-C case definition which has been in place since May 2020 that patients must fulfill for reporting cases.^5^

The onset of symptoms in children with MIS-C occurs 2-6 weeks after symptomatic or asymptomatic SARS-CoV-2 infection. Current CDC criteria includes a diverse array of organ-specific manifestations and laboratory findings; a final criterion is the need for inpatient admission. With this requirement for inpatient admission, potentially milder forms of MIS-C that were managed in the outpatient setting have not been appreciated. The existence of such cases would ultimately necessitate a revision of the current case definition. Previous studies of MIS-C originated in intensive care settings or relied on existing inpatient networks repurposed from other disease conditions, so identifying outpatient cases would fall outside the designed infrastructure.^6-10^ Using the electronic health record (EHR) data from the PEDSnet clinical research network, we sought to identify and describe the existence of an outpatient cohort of children with MIS-C.

## Methods

PEDSnet (pedsnet.org) is a national clinical research network of children’s health systems that share standardized data drawn from electronic health records (EHR) as part of collaborative research to improve child health^11^. Participating institutions included Children’s Hospital of Philadelphia, Cincinnati Children’s Hospital Medical Center, Children’s Hospital Colorado, Ann & Robert H. Lurie Children’s Hospital of Chicago, Nationwide Children’s Hospital, Nemours Children’s Health System (a Delaware and Florida health system), Seattle Children’s Hospital and Stanford Children’s Health. The PEDSnet COVID-19 Database Version 2022-06-06 was used for this study. The Children’s Hospital of Philadelphia’s institutional review board designated this study as not human subjects research and waived the need for consent.

Patients were identified via two distinct methods, both of which derive from the treating clinician’s diagnosis. The first is the use of an MIS-C ICD-10CM code (M35.81). The other is a string searching method evaluating matches in the diagnosis term from the proprietary point-of-care terminology (Intelligent Medical Objects (Rosemont, IL). A match is made via string searching when the diagnosis term contains the token ‘MIS-C’ or when both the token ‘multisystem’ and the subword token ‘inflam’ are present in the same term. For patients so identified, information about encounters, including setting (inpatient, outpatient, emergency department), diagnoses, medications, and laboratory test results related to MIS-C diagnostic criteria were retrieved from the PEDSnet database. A group of MIS-C cases adjudicated by individuals at each institution for the purposes of public health reporting using CDC criteria was included for comparison.

All laboratory results were standardized to common units to allow for comparison across health systems, and a set of thresholds for abnormal values consistent with the CDC definition for MIS-C was used. Descriptive statistics were computed using R, version 3.2.0 (Vienna, Austria)^12^.

## Results

Across all institutions, we identified 1569 children with MIS-C, and of these 209 did not have an associated inpatient admission within 90 days of their MIS-C diagnosis. Due to the possibility that many of these children were admitted to other institutions and were being evaluated during follow-up outpatient visits, we conducted systematic manual chart reviews for all 209 patients. Ultimately, we identified 7 children from 2 of the 9 participant institutions who were managed without inpatient admission with a physician-assigned diagnosis of MIS-C. Table 1 is a summary of the demographic and clinical information on these 7 patients compared to a cohort of inpatients with MIS-C who were reported to public health institutions as confirmed cases. These children were all male (100%) and mostly younger than age 11 years (54%). They were all seen in the Emergency Department for signs and symptoms characteristic of MIS-C: fever (100%), abdominal pain (85%), headache (57%), rash (43%), nausea/vomiting (29%) and mucosal involvement (29%). These children did have laboratory results consistent with those observed in MIS-C including elevated C-reactive protein (CRP), lymphopenia, elevated troponin-I or T, elevated ferritin, elevated d-dimer and elevated creatinine. Figure 1 summarizes the laboratory values compared to inpatients with confirmed MIS-C and illustrates how this cohort of patients had some results that clearly crossed outside the normal range of values. These children were treated with intravenous fluid resuscitation, responded well and were discharged home for possible follow up as outpatients. Two received oral corticosteroids and none received aspirin or other immunomodulators. One patient had an observation status admission but did not receive any MIS-C related treatment. Review of follow-up visits for all 7 patients did not demonstrate subsequent admissions, or complications related to MIS-C or PASC.

**Table 1.**
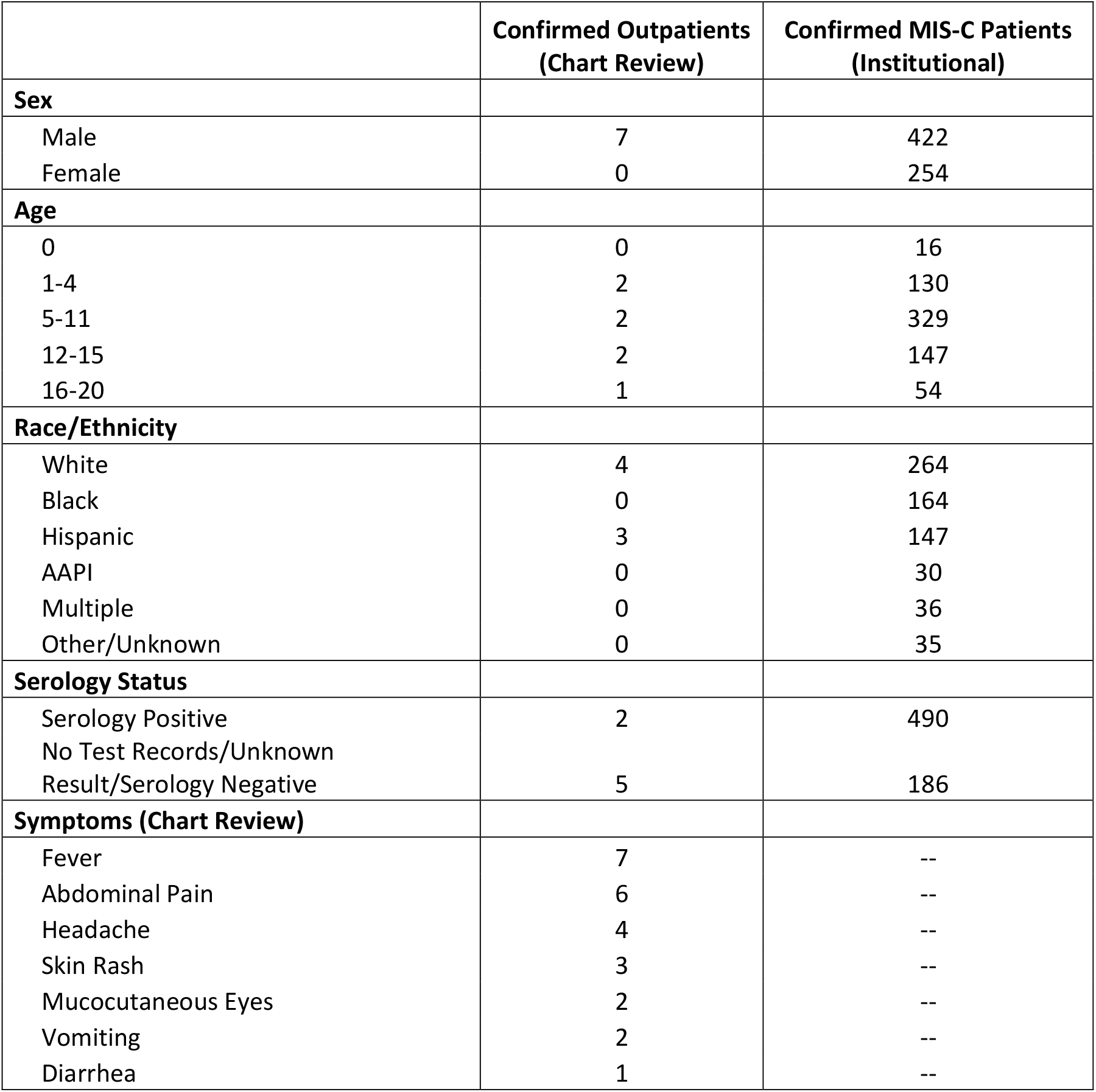
Demographic and Clinical Summary of Children with MIS-C Managed as Outpatients.

|  | **Confirmed Outpatients**<br>**(Chart Review)** | **Confirmed MIS-C Patients**<br>**(Institutional)** |
| --- | --- | --- |
| **Sex** |  |  |
| Male | 7 | 422 |
| Female | 0 | 254 |
| **Age** |  |  |
| 0 | 0 | 16 |
| 1-4 | 2 | 130 |
| 5-11 | 2 | 329 |
| 12-15 | 2 | 147 |
| 16-20 | 1 | 54 |
| **Race/Ethnicity** |  |  |
| White | 4 | 264 |
| Black | 0 | 164 |
| Hispanic | 3 | 147 |
| AAPI | 0 | 30 |
| Multiple | 0 | 36 |
| Other/Unknown | 0 | 35 |
| **Serology Status** |  |  |
| Serology Positive | 2 | 490 |
| No Test Records/Unknown<br>Result/Serology Negative | 5 | 186 |
| **Symptoms (Chart Review)** |  |  |
| Fever | 7 | -- |
| Abdominal Pain | 6 | -- |
| Headache | 4 | -- |
| Skin Rash | 3 | -- |
| Mucocutaneous Eyes | 2 | -- |
| Vomiting | 2 | -- |
| Diarrhea | 1 | -- |

**Figure 1.**
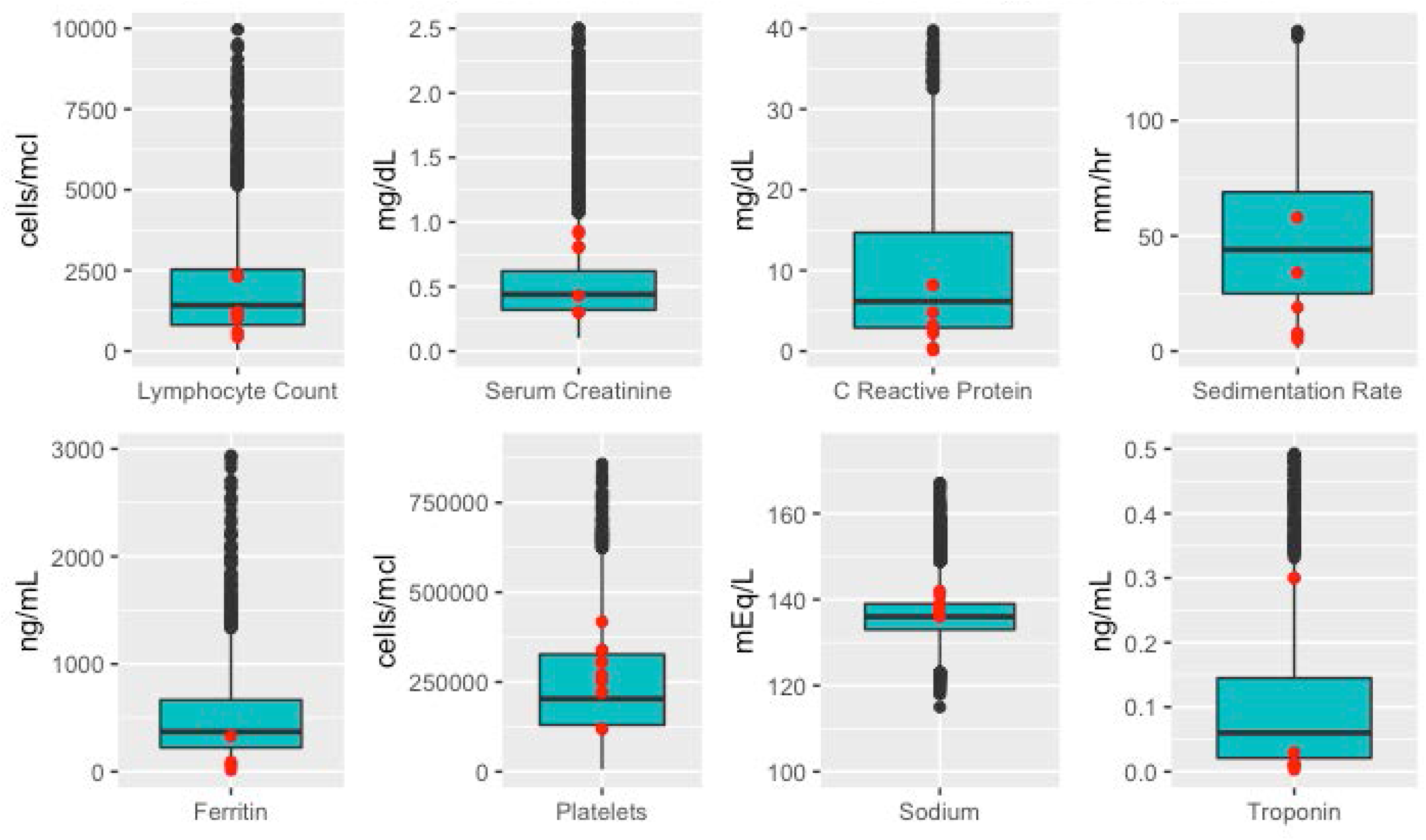
Laboratory Results of Children with MIS-C Managed as Outpatients. For each displayed laboratory test, the distribution of the resultant measurements for both MIS-C outpatients and confirmed MIS-C inpatients is represented in a boxplot (blue) bounded at the 25^th^ and 75^th^ percentiles, while the distribution for the outpatient population is overlayed in red points. All lab results are included in each distribution, so a patient with two serum creatinine measures would contribute two measurements.

## Discussion

Using the PEDSnet data infrastructure, we identified a subset of outpatient children with a mild form of MIS-C who were managed without the need for hospitalization and largely without systemic immunomodulatory treatment. To our knowledge, this is the first description of exclusively outpatient MIS-C. Because PEDSnet collects data from member children’s hospitals that encompass both inpatient and outpatient settings, we are in a unique position to describe this rare cohort.

These children had laboratory abnormalities consistent with those children meeting criteria for MIS-C based on the CDC case definition, but the degree of these abnormalities was not as severe. These children were treated with fluid resuscitation in the emergency department with only a minority who received oral steroids. We did not identify any prior or subsequent admissions for any of these children, which suggests that this mild phenotype remained consistent throughout their course of illness and appears to be associated with good outcomes.

While this is a small cohort of children managed as outpatients, it is important to acknowledge how institutional practices influence the management of MIS-C. These 7 children were identified in only 2 institutions, and several PEDSnet institutions recommended inpatient management for any child in whom the diagnosis of MIS-C was being considered. It is unclear if this was due to an abundance of caution managing a novel condition, or whether inclusion of inpatient admission in the CDC case definition influenced practices. Across 5 of our PEDSnet institutions, there were 47 children out of the 1569 total who were admitted with MIS-C, improved rapidly and were discharged home the next day. it is tempting to speculate that these patients represent a similar phenotype that could be managed expectantly as outpatients. Of note, our network has analyzed this same EHR data using latent class analysis to identify a cohort of patients with a mild phenotype of MIS-C that is distinct from the more severe presentations that have been traditionally associated with MIS-C. It is likely that MIS-C comprises a wider illness spectrum, and these 2 cohorts represent a proportion of children with mild MIS-C who may not require the same treatment regimen as children with more severe presentations and who may improve with supportive care and observation alone.

There are some limitations to acknowledge with these results. PEDSnet institutions comprise open integrated delivery systems where patients use services at their discretion. This can create gaps in EHR data capture, and our chart review demonstrated this. We found that 202 of the 209 patients with no inpatient encounters in the PEDSnet database who were referred for outpatient specialty care management following hospitalization elsewhere. Given the tertiary/quaternary status of the children’s hospitals within PEDSnet, it is unlikely that the children in our cohort evaluated and managed in the ED sought admission elsewhere. This was a retrospective case series, so diagnosis of MIS-C was based on clinical judgement with variability from person to person and site to site. It is possible these children had another diagnosis with features overlapping with MIS-C. Given that each child had a prior COVID-19 infection or high-risk exposure and most had serologic confirmation, we think this is unlikely. We acknowledge this is a small group of patients. Finally, it is possible that we have underestimated the number of children with MIS-C treated in outpatient settings because some physicians did not assign this diagnosis since the patient was not admitted and did not meet CDC criteria.

These findings should be considered a discovery cohort and should spur further investigation. Systematic review of other appropriate data sources and prospective studies with adequate follow-up should be conducted to identify children diagnosed with MIS-C who are managed as outpatients in a similar manner to the patients identified in our cohort. If other studies corroborate these findings, an update of the CDC definition may be required to remove the requirement of inpatient admission for the confirmation of an MIS-C diagnosis. In conclusion, our cohort of children with mild presentations of MIS-C, managed without anti-inflammatory medications, suggest that inpatient admission and directed treatment may not be required for all patients, with potential to reduce unnecessary health care utilization, while maintaining good long-term outcomes.

## Data Availability

All data produced in the present study are available upon reasonable request to the authors.

## Acknowledgements

This study is part of the NIH Researching COVID to Enhance Recovery (RECOVER) Initiative, which seeks to understand, treat, and prevent the post-acute sequelae of SARS-CoV-2 infection (PASC). For more information on RECOVER, visit https://recovercovid.org/.

The authors wish to thank Jordan Musante and Miranda Higginbotham for their assistance with project management, as well as colleagues across the PEDSnet sites who helped support the data infrastructure used for this work. We would also like to thank the National Community Engagement Group (NCEG), all patient, caregiver and community Representatives, and all the participants enrolled in the RECOVER Initiative.

## Notes

Conflicts of Interest: Dr. Jhaveri is a consultant for AstraZeneca, Seqirus and Dynavax, and receives an editorial stipend from Elsevier. Dr. Mejias reports funding from Janssen, Merck for research support, and Janssen, Merck and Sanofi-Pasteur for Advisory Board participation; Dr. Schulert reports consulting fees from Novartis and SOBI; Dr. Rao reports prior grant support from GSK and Biofire and is a consultant for Seqirus.

### Competing Interest Statement

Dr. Jhaveri is a consultant for AstraZeneca, Seqirus and Dynavax, and receives an editorial stipend from Elsevier. Dr. Mejias reports funding from Janssen, Merck for research support, and
Janssen, Merck and Sanofi-Pasteur for Advisory Board participation; Dr. Schulert reports consulting fees from Novartis and SOBI; Dr. Rao reports prior grant support from GSK and Biofire and is a consultant for Seqirus.

### Funding Statement

This research was funded by the National Institutes of Health (NIH) Agreement OT2HL161847-01 as part of the Researching COVID to Enhance Recovery (RECOVER) program of research.

### Author Declarations

The Children's Hospital of Philadelphia's institutional review board designated this study as not human subjects research and waived the need for consent.

